# What influences non-menstruator attitudes and behaviours towards menstruation among Rohingya refugees in Bangladesh? A quantitative analysis

**DOI:** 10.1101/2024.09.17.24313147

**Authors:** Georgia Hales, Paul Hutchings, Katy Roelich, Mahua Das, Nowshad Akram, Shajeda Begum, Zahida Sultana

## Abstract

Non-menstruators play an important yet overlooked role in shaping menstrual health. They may be the family budget holders who purchase menstrual materials, receive health information outside the household, or preserve social stigmas. In response, World Vision, supported by UNICEF, implemented a programme to influence non-menstruators among the Rohingya population living in Kutupalong refugee camp, Bangladesh. The intention was to ensure non-menstruators recognise their important role in supporting family members to improve their menstrual health. We evaluated this intervention using a baseline and endline survey of 150 participants based on the Risk-Attitudes-Norms-Ability-Self-regulation (RANAS) approach to behaviour change. Based on the responses, we categorised participants into ‘doers’ and ‘non-doers’ of positive behaviours. We then performed multiple linear regression analyses and ANOVA comparison of means tests to understand how psychosocial and contextual factors influence the population’s attitudes and behaviours towards menstruation before and after intervention. The regression analyses showed eight psychosocial and contextual factors that had a significant relationship with the desired behaviours. These were the block they lived in, marital status, already having learnt about menstruation from World Vision, how they were introduced to menstruation, who they discuss menstruation with, family members’ reaction, perceptions of their role, and their commitment. Additionally, the ANOVA comparison of means between doers and non-doers showed doers were more likely to be confident to carry out the behaviour, perceive their role as important, and discuss menstruation with family. They were also more likely to be married, older, and have first learnt about menstruation from their family. This is the first study to identify which psychosocial and contextual factors significantly influence positive non-menstruator attitudes and conducive behaviours towards menstruation and menstruators. Humanitarian organisations can use these factors to improve the design and targeting of behaviour change programmes to improve menstrual health.

**Highlights:**

1. Menstrual health is often limited in refugee settings
2. Non-menstruators can enable and constrain menstrual health
3. Menstrual health intervention among Rohingya refugees shows positive change in non-menstruator behaviour
4. Behavioural determinants for non-menstruators related to marriage, commitment, and family reaction
5. ‘Doers’ more likely to be confident, perceive role as important, and discuss menstruation with family

## Introduction

Only in the last 10 to 15 years have we started to consider menstrual health as a public health and human rights issue and consequently began to address it as such (Babbar et al. 2022). Even more recently (in 2021) have we developed a definition for menstrual health. The definition highlights how it is more than just a ‘women’s issue’ in more ways than one, emphasising the need to expand our understanding of gender beyond the binary in that it is not only cis-gender women who menstruate but also transgender men and other gender diverse persons. Accordingly, we use the terms ‘menstruator’ to denote those who have the ability to menstruate and ‘non-menstruator’ for those who do not.

Though inadequate menstrual health is a global issue, it is exacerbated in certain contexts. In the humanitarian sector, for example, it has been a neglected topic. This has resulted in a failure to meet the basic human needs among some of the most marginalised groups in the world. It can be ascertained that this is due to the stigmatised and secretive nature of menstruation, a disregard for issues perceived to concern only women, and a lack of coordination; since menstrual health falls into multiple departments within a humanitarian response, no sector was taking responsibility for it (Sommer et al. 2016). It was not until 2000 when the UNFPA recommended that ‘dignity kits’ with menstrual materials be part of the reproductive health response (Tellier et al. 2020). However, these were often largely insufficient (Rohwerder 2014). Very gradually the scope widened beyond materials where – 16 years later - Sommer et al. (2016) proposed that the response also requires facilities for washing, changing, drying, and disposing of materials alongside supplying menstrual education and practical information. Following this, the MHM in Emergencies Toolkit was developed in 2017 and both hardware and software MHM guidelines were written into the 2018 Sphere Standards for humanitarian action. Only now are we talking about applying a holistic approach where we address the root causes of inadequate menstrual health – menstrual taboo – and involve all members of society in this endeavour, not just those who menstruate.

These standards and guidelines have supported interventions that have improved menstrual health alongside reducing menstrual stigma. Since menstruation is often private and taboo, interventions have logically focused only on women, or those who menstruate. However, researchers and practitioners have come to observe that non-menstruators can be an impediment to progress (Day 2024; Patel et al. 2022). All members of a society can hold misinformed and stigmatised views on menstruation, however the portion of the population who have been excluded are typically the ones to hold more power and influence. To illustrate, non-menstruators can perpetuate negative stigmas and taboos about menstruation that reinforce beliefs and practises that shame and marginalise menstruating individuals. This stigma often leads to secrecy and a lack of open discussion, which in turn means menstrual health issues are not well understood or prioritised. It also endorses misogyny and resultantly upholds gender inequalities.

In the last couple of years, some humanitarian organisations have started to recognise the importance of including non-menstruators in menstrual health programmes. Due to its infancy, there is little documentation of these approaches nor research into their effectiveness. This means there is a lack of research-based guidance for humanitarian organisations addressing this facet of menstrual health. What is also lacking is an exploration of why these negative or avoidant attitudes and behaviours towards menstruation endure.

To address this, we studied a World Vision and UNICEF behaviour change programme with non-menstruator Rohingya refugees living in Kutupalong camp, Bangladesh, focused on reducing stigma around menstruation and encouraging supportive behaviours towards their menstruating family members. We conducted a longitudinal baseline and endline survey of 150 participants using the Risk-Attitudes-Norms-Ability-Self-Regulation (RANAS, 2022) framework, which we analysed using multiple linear regression and ANOVA comparison of means tests. The aim was to understand which contextual and psychosocial factors influence positive attitudes and conducive behaviours towards menstruation among this population. To do this we addressed two research questions: (RQ1) which contextual and psychosocial factors are behavioural determinants for having positive attitudes and conducive behaviours towards menstruation and menstruators? And (RQ2) which psychosocial factors have changed significantly after World Vision’s engagement of Rohingya non-menstruators in a menstrual health programme?

## Methods

### Behaviour change approach and theory

The RANAS approach provides a comprehensive framework for understanding what influences behaviours. It is based on empirical evidence and behavioural science theories drawing on psychology and sociology to identify key determinants of behaviour. It works by discovering the ‘doers’ and ‘non-doers’ of a behaviour, analysing differences in their psychosocial orientations, and using this to select Behaviour Change Techniques (BCTs), tailored to the specific context and population. The approach - developed in 2012 – has been used to evaluate and develop water, sanitation, and hygiene (WaSH) behaviour change programmes and has been used before in the same setting as the one studied in this paper (Rahamen 2022). The research partners (World Vision and UNICEF) also use this approach in their programming. It is more relevant than other behaviour change frameworks as it is tailored to the population, has proven to effectively change behaviour under local conditions, saves resources due to adapted interventions, and provides an evidence base for further interventions and upscaling (RANAS 2023).

### Study population and design

The Rohingya refugee population living in Bangladesh due to persecution from Myanmar are a patriarchal community. Their social structures – combined with the challenges of living in a refugee camp - impinge on menstruators’ menstrual health on multiple levels: materials and ability for washing, changing, drying, and disposing of them, safety, stigmatisation, a supportive environment, and access to education, knowledge, healthcare, and overall health (Pandit et al. 2022). This paper does not intend to criticise or single-out this population but rather recognise that menstruation is stigmatised globally and that non-menstruators can affect menstrual health in any context, though its manifestations vary (Winkler et al., 2024). They provide a good example of a case where the social structures and deep-rooted beliefs about menstruation impinge greatly on the population’s menstrual health. It is also is one of the few places where humanitarian organisations are addressing the issue. The study included 150 non-menstruators in a UNICEF-run camp within the wider Kutupalong refugee camp, Cox’s Bazar, Bangladesh. We carried out a longitudinal study using a survey either side of a World Vision intervention at baseline (April 2023) and endline (March 2024). The World Vision data collection team conducted face-to-face surveys among the six blocks within the camp.

### Sampling

Since there are no studies on quantitative evidence for changing menstrual attitudes and behaviours among non-menstruators, we undertook two statistical power calculations based on results from of a paper measuring multiple social norms and beliefs regarding gender based violence (GBV) before and after intervention (Glass et al. 2019). One was for beliefs based on a husband’s right to use violence, the other regarded protecting family honour. From the data, the following means and the standard deviations were calculated. Using a sample size calculator (Power and Sample Size, 2024), the researchers calculated the required number of participants based on a 2-sample, 1 sided power sample to Power 1-β : 0.80; Type I error rate: 5%; Sampling ratio: 1. The sample sizes were 205 and 53 respectively, the mean of which is 129. Thus, we used a sample size of 150 to provide a small error margin as some surveys may have errors. For the baseline survey only 149 out of the target sample of 150 people were available. For the endline survey 146 were available. They were not replaced with substitutes. Participants were randomly selected from resident lists including a proportionate number of people depending on the number of households within each sub-block.

**Table 1.**
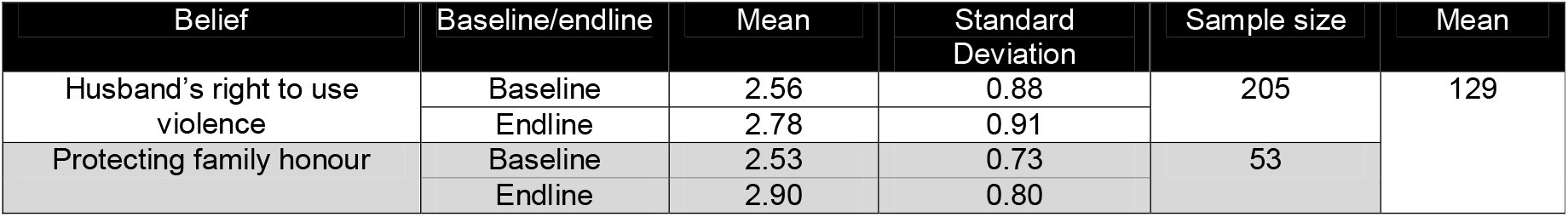
Sample size calculation using two statistical power calculations.

### Data collection methods and training

Nine data collectors gathered quantitative data on a shared Excel spreadsheet using the same survey questions at baseline and endline. Prior to data collection, the lead author met with the World Vision research partners and data collection team to appraise survey questions, discuss the procedure, and communicate the goals and theoretical background of the survey. As a pilot, the lead author conducted the first 10 surveys with one World Vision research partner (an author of this paper) and one member of the data collection team to verify its applicability. From this, we removed some less-relevant questions to reduce participants’ time. Data collectors had prior interview experience and rapport with the community. The participants had been briefed about the nature of the survey before participating. The research partners coordinated and monitored the survey procedure throughout the period of data collection.

### Questionnaires and measures

The structured, face-to-face survey questions and responses were written in English but conducted in Rohingya – a spoken language without a written form, which is closely related to the Bengali dialect of Chittagong with some accent and pronunciation differences – in which the data collectors are native. The researchers designed the questionnaire using the psychosocial factors from the RANAS model, alongside contextual and behavioural questions based on literature on non-menstruator Rohingya living in Kutupalong camp. Some questions were closed on Likert scales. Those that were open were later categorised and quantified on scales to give participants more freedom to answer in a way they deemed suitable. As an example, non-menstruators gave responses to the question ‘When someone menstruates what do you think of them?’ ranging from ‘no idea’ to ‘It’s a natural issue for women and during this time family member should support them both mentally and physically’. We then created a scale that fitted the responses from negative, neutral to positive. The survey covered contextual factors, behaviours, health awareness and risk, attitudes, norms, confidence, and commitment. Contextual factors included area of residence within the camp, age, education level of self, father and mother, marital status, parental status, and arrival date in camp.

### Statistical analysis of data

We performed statistical analyses of frequencies, correlations, ANOVAs (Analysis of Variance), and multiple linear regressions in Microsoft Excel 2016 using the ‘Regression’ and ‘ANOVA: Single Factor’ plug-ins. For the two regression analyses we used support offered to menstruating family members, feelings towards supporting family members and attitudes towards menstruation as a combined outcome (dependent variable) and (1) contextual factors and (2) psychosocial factors of the RANAS model as predictors (independent variables). Multiple linear regression is a statistical technique used to analyse the relationship between two or more independent variables and a dependent variable. It determines which factors have the most significant impact on the outcome. We also conducted two ANOVA means test: one for ‘doers’ and ‘non-doers’ (of the outcome behaviour) and the other for baseline and endline results to determine statistically significant differences.

### Ethics

Ethical approval was gained from the University of Leeds on the 16^th^ March 2023 under reference code MEEC 22-019.Lead author access to Kutupalong Camp was granted by The Government of Bangladesh’s Refugee Relief and Repatriation Commissioner on the 21^st^ March 2023. The lead author provided an explanatory statement, consent, and confidentiality form to the World Vision data collection team. Since Rohingya is a spoken, rather than written language, the data collection team verbally translated it to participants upon recruitment and took verbal consent. There was always more than one data collector present. Recruitment occurred from April 17-30^th^ 2023.

## Results

### Demographics

Below is the demographical data of study participants at baseline. Religion is excluded as a demographical category as all participants were Muslim. The majority of Rohingya arrived in Bangladesh in 2017 after a massive wave of violence broke out where they resided in Rakhine State, though their persecution started before this in the 1990s. Hence we categorise the arrival to the camp as before, during and after the 2017 influx. They have been divided into doers and non-doers following the RANAS approach. Non-doers were those who answered negatively to at least one of the following three questions:

- What support do you offer when a family member is menstruating?
- How do you feel to support your family members during their menstruation?
- When people menstruate what do you think of them?

**Table 2.**
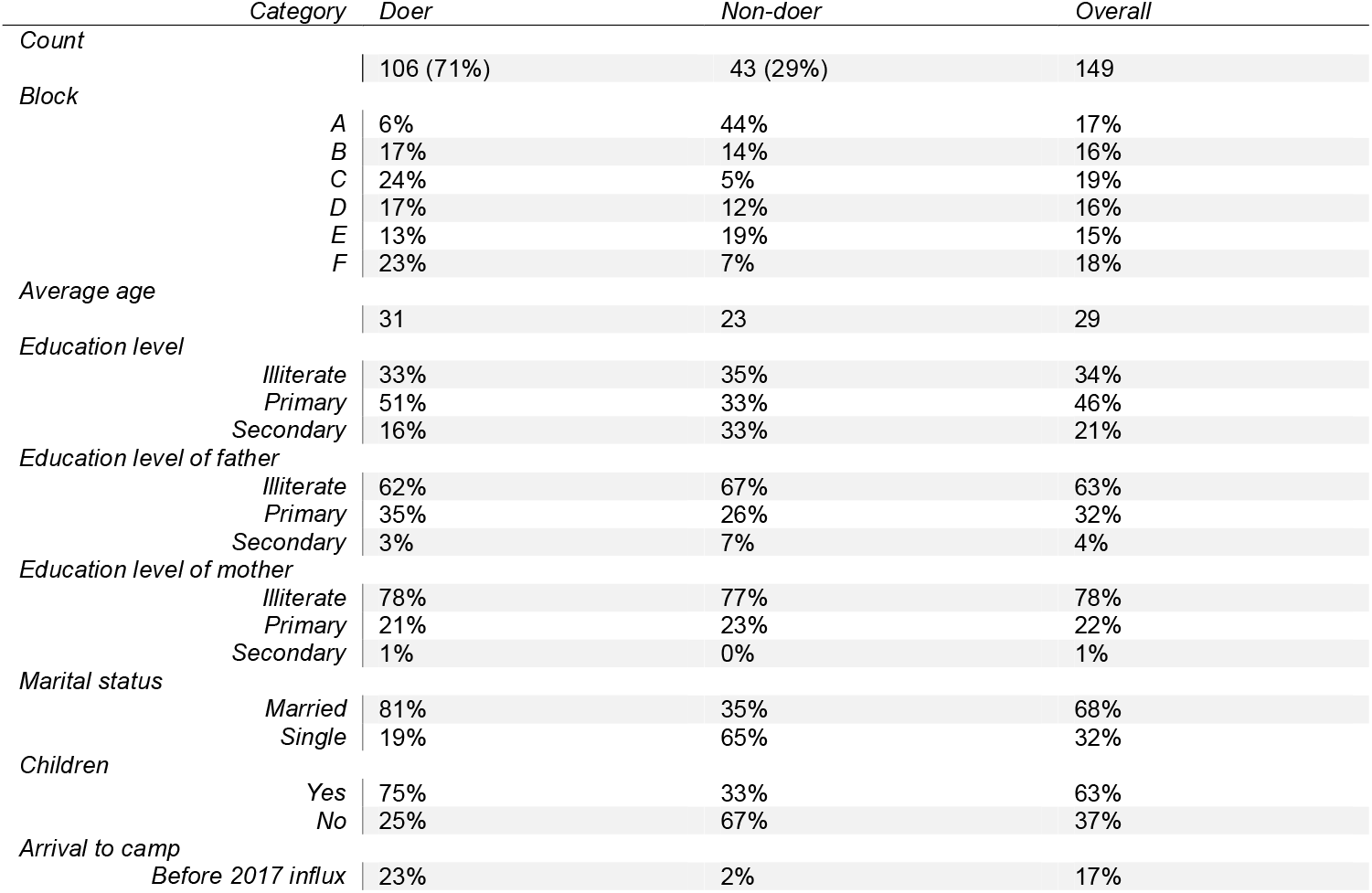
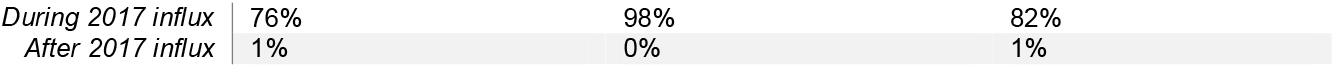
Demographics – baseline doers, non-doers and overall.

|  | Category | Doer | Non-doer | Overall |
| --- | --- | --- | --- | --- |
| Count |  | 106 (71%) | 43 (29%) | 149 |
| Block |  |  |  |  |
|  | A | 6% | 44% | 17% |
|  | B | 17% | 14% | 16% |
|  | C | 24% | 5% | 19% |
|  | D | 17% | 12% | 16% |
|  | E | 13% | 19% | 15% |
|  | F | 23% | 7% | 18% |
| Average age |  | 31 | 23 | 29 |
| Education level |  |  |  |  |
|  | Illiterate | 33% | 35% | 34% |
|  | Primary | 51% | 33% | 46% |
|  | Secondary | 16% | 33% | 21% |
| Education level of father |  |  |  |  |
|  | Illiterate | 62% | 67% | 63% |
|  | Primary | 35% | 26% | 32% |
|  | Secondary | 3% | 7% | 4% |
| Education level of mother |  |  |  |  |
|  | Illiterate | 78% | 77% | 78% |
|  | Primary | 21% | 23% | 22% |
|  | Secondary | 1% | 0% | 1% |
| Marital status |  |  |  |  |
|  | Married | 81% | 35% | 68% |
|  | Single | 19% | 65% | 32% |
| Children |  |  |  |  |
|  | Yes | 75% | 33% | 63% |
|  | No | 25% | 67% | 37% |
| Arrival to camp |  |  |  |  |
|  | Before 2017 influx | 23% | 2% | 17% |

|  |  |  |  |
| --- | --- | --- | --- |
| During 2017 influx | 76% | 98% | 82% |
| After 2017 influx | 1% | 0% | 1% |

### Contextual behavioural determinants

To answer the first research question, the researchers identified which contextual factors influence positive menstrual attitudes and conducive behaviours through applying a multiple linear regression analysis using self-reported attitudes and behaviours as the outcome and contextual factors as predictors. All study participants from the baseline level were included in the analysis. This model explained 48.3% of the variance in the menstrual attitudes and behaviours. Multicollinearity reduces the precision of estimate coefficients, which weakens the statistical power of a regression model. Therefore, because marital status and having children was highly correlated (89%), having children was excluded from the regression analysis. Those factors with a correlation of 20% or above with positive menstrual attitudes and conducive behaviours were included in the analysis. See supplementary materials for the factors, which were excluded.

Four factors were significant predictors of having positive menstrual attitudes and conducive behaviours (see these factors highlighted in **Error! Reference source not found**.). They were: location within the camp (living in Block A [β = -1.127], living in Block F [β = 1.062]), being married (β = 1.104), already having learnt about menstruation from World Vision (β = 0.552), and having first learnt about menstruation from an Imam or the Quran (β = -1.094). This means that an increase in positive attitudes and conducive behaviours can be expected if one lives in Block F, is married, or has already learnt about menstruation from World Vision, if all other factors remain the same. And that a decrease in positive attitudes and behaviours can be expected if one lives in Block A, or first learnt about menstruation from the Quran, if all other factors remain the same. The Beta value indicates the percentage change to be expected in the desired behaviour with every 1-point increase of the independent variable. Here we show the percentage increase from respondents who gave a score of zero compared with the top number of the scale. An increase in positive attitudes and conducive behaviours of 106% can be expected for those living in Block F compared to those who do not. These attitudes and behaviours can also be expected to increase by 110% in those who are married, and by 55% in those who have already learnt about menstruation from World Vision. Those living in Block A are expected to have a decrease in positive attitudes and conducive behaviours of 113%, along with a decrease of 109% for those who first learnt about menstruation from an Imam or the Quran. Looking at the contextual factors presents ideas to either target or be aware of within behaviour change interventions. Those factors, which cannot be changed, for example marital status, are more to inform why certain groups may hold certain opinions or perform certain actions. It may indicate areas where different groups need to be targeted in different ways.

**Table 3.** Multiple linear regression – contextual factors - Adjusted R^2^ = 0.483 N=149, highlighted blue = p<0.05. All factors are on a Yes/No =1/0 scale apart from age which is on an interval scale.

| Contextual factor | M(SD) | $\beta$ | p-value |
| --- | --- | --- | --- |
| Lives in Block A | 0.17 (0.37) | -1.127 | 0.0001 |
| Lives in Block C | 0.18 (0.39) | 0.112 | 0.8001 |
| Lives in Block F | 0.18 (0.39) | 1.062 | 0.0003 |
| Age | 29 (9) | -0.001 | 0.9293 |
| Marital Status | 0.68 (0.47) | 1.104 | 0.0001 |
| Arrived during 2017 influx | 0.83 (0.38) | -1.679 | 0.1337 |
| Arrived before 2017 influx | 0.17 (0.37) | -1.445 | 0.2198 |
| Already learnt about menstruation from WV | 0.49 (0.50) | 0.552 | 0.0093 |
| Learnt about menstruation age 5-10 | 0.05 (0.23) | -0.313 | 0.4744 |
| Learnt about menstruation age 15-20 | 0.40 (0.49) | -0.305 | 0.1537 |
| Learnt about menstruation age 21-30 | 0.21 (0.41) | -0.024 | 0.9398 |
| First learnt about menstruation from family | 0.39 (0.49) | -0.408 | 0.1098 |
| First learnt about menstruation from Imam/Quran | 0.04 (0.20) | -1.094 | 0.0200 |
| First learnt about menstruation from NGO | 0.07 (0.25) | -0.252 | 0.5479 |

### Psychosocial behavioural determinants

The researchers used the same method for the psychosocial factors. The model explained 59.5% of the variance in the menstrual attitudes and behaviours. Since there were many factors included in the analysis, those with a correlation of 20% or above with the dependent variable were included in the analysis. See supplementary material for a list of these factors.

Four factors were significant predictors of having positive menstrual attitudes and conducive behaviours (see the factors highlighted in Table 4). They were: discussing menstruation with no one (β = -0.502), feeling that menstruating family members would feel positive if they supported them during menstruation (β= 0.547), considering their role to be important (β= 0.447), and feeling committed to supporting their family with their menstrual health (β= 0.370). This means that an increase in positive attitudes and conducive behaviours can be expected if any of these four significant factors increases or decreases while all other factors remain the same. So a decrease in positive attitudes and conducive behaviours of 50% can be expected in respondents who do not discuss menstruation with anyone. Positive attitudes and conducive behaviours are expected to increase by 110% in respondents who believe their menstruating family members will feel happy that they support them during menstruation compared to those that don’t. An increase of 90% can be expected in respondents who perceive their role in supporting family members with their menstrual health is very important, and by 74% in respondents who feel strongly committed to supporting their family with their menstrual health. Accordingly, if we target these significant psychosocial factors with specific behaviour change interventions, we can expect people to be more likely to acquire more positive attitudes and conducive behaviours towards menstruation and menstruating family members.

**Table 4.**
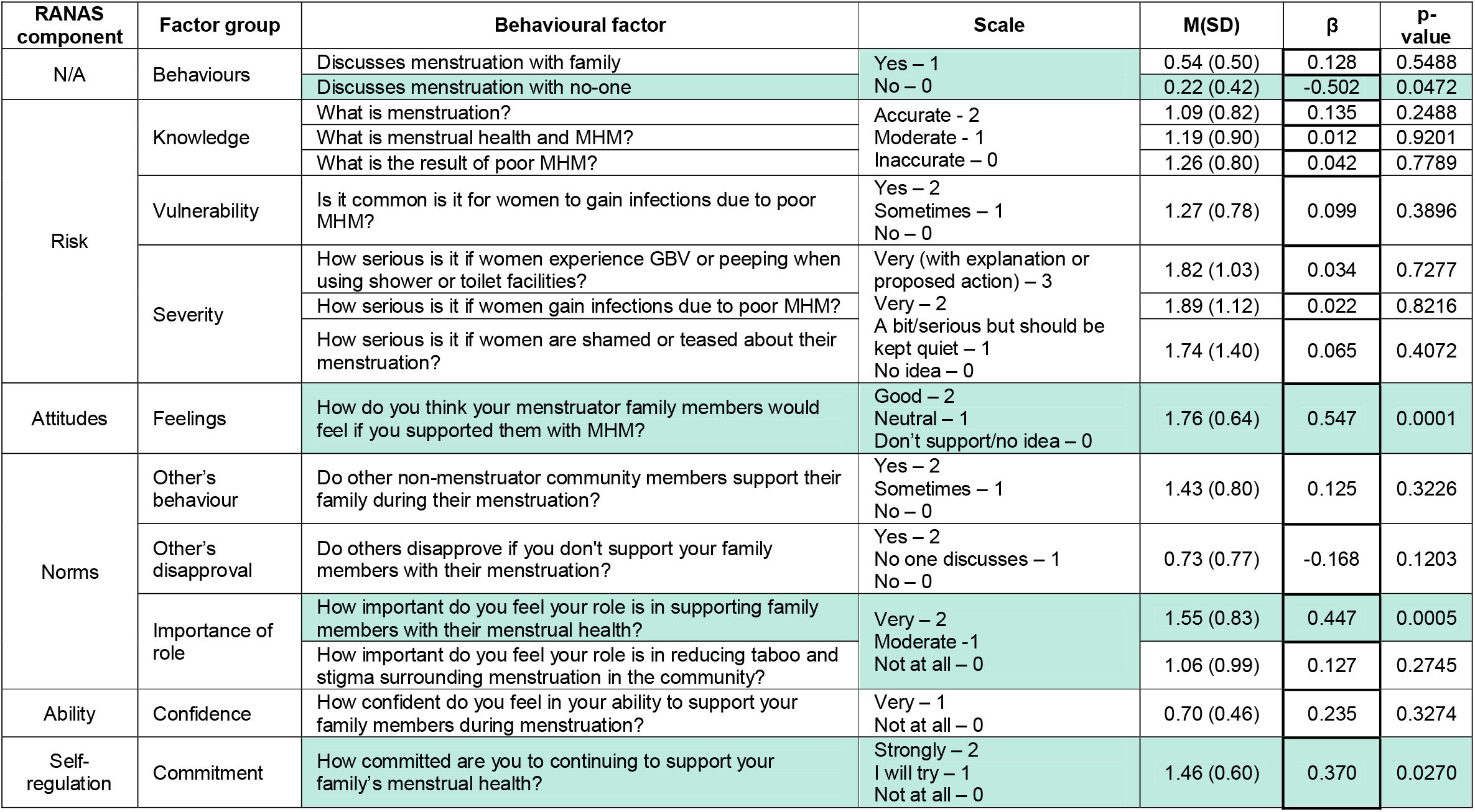
Multiple linear regression – RANAS psychosocial factors - Adjusted R^2^ = 0.595, N=149, highlighted blue = p ≤0.05.

### Doer/non-doer ANOVA analysis contextual factors

To further explore which contextual factors are important to consider or alter, the researchers undertook a doer/non-doer analysis following the RANAS approach. The researchers used ANOVA comparison of means to understand which response averages had a significant difference between those who did and did not do the behaviour. Only responses with a p-value ≤_J.05 are shown in **Error! Reference source not found**.. The scales for each category were binary (Yes/No) apart from age. To make them cohesive, age has been put on a binary scale of ‘over/ under the age of 25’. **Error! Reference source not found**. then shows what percentage of people answered ‘yes’ to the categories within the doer and non-doer groups. The results are in order of biggest negative to positive difference between doers and non-doers.

Non-doers were more likely to live in block A and doers in block C and F, coinciding with the regression analysis. They were also more likely to have arrived during the 2017 influx, learnt about menstruation age 5-20, and learnt about menstruation from friends or the Imam/Quran. Doers were more likely to be age 25 or older, married, have children, have previously learned about menstruation from World Vision and to agree to take part in World Vision’s next menstrual health campaign. Doers were more likely to have arrived before the 2017 influx, have first learned about menstruation age 21-30, and first learned from family and NGOs.

**Figure 1.**
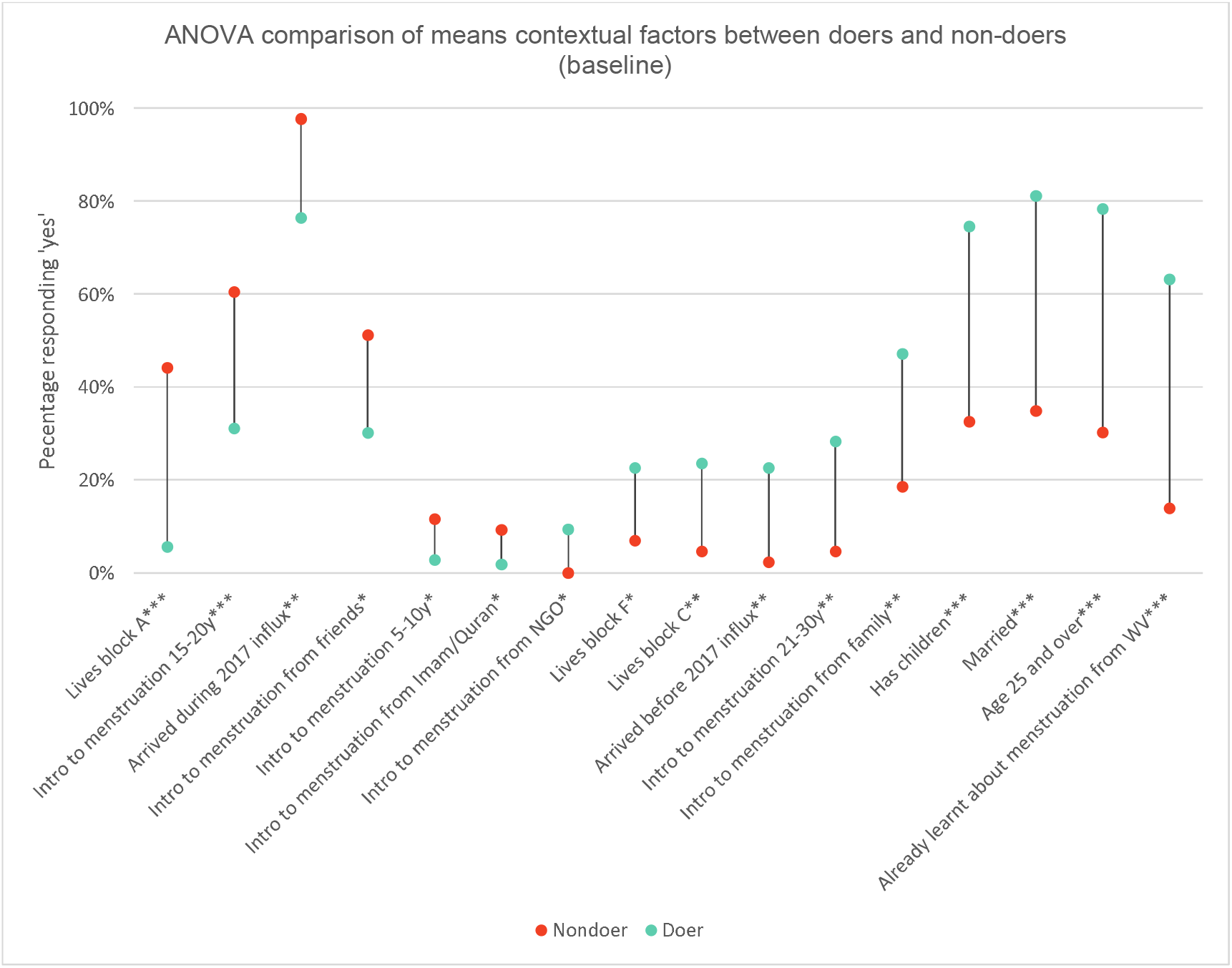
ANOVA comparison of means for contextual factors for doers and non-doers - ^*^p□≤□.05, ^**^p□≤□.01, ^***^p□≤□.001. n = 149

### Doer/non-doer ANOVA analysis psychosocial factors

The researchers undertook the same method for the psychosocial factors. Only responses with a p-value ≤_J.05 are shown in **Error! Reference source not found**.. The scales were the same as for the multiple linear regression for RANAS psychosocial factors yet they have all been put on the same scale from ‘strongly disagree’ to ‘strongly agree’ to make them visually comparable. The results are in order of biggest negative to positive difference between doers and non-doers.

Doers were more likely to believe that it is a serious matter for menstruators not to participate in normal daily activities, experience GBV or peeping when using WaSH facilities, or gain infections due to poor MHM. Doers were also more likely to believe it was acceptable for menstruators to cook food during menstruation. Doers reported feeling that their role in supporting family with their menstrual health and reducing menstrual stigma in the community was more important than non-doers. Doers were more likely to discuss menstruation with their family, whereas non-doers were more likely to discuss menstruation with their (non-menstruator) friends or no one at all. As expected, doers gave more accurate descriptions of menstruation, menstrual health, MHM, and the results of poor MHM. Doers also reported feeling more confident and committed to supporting their menstruating family. In terms of perceptions of the actions of other community members, doers believed that other community members also support their family during menstruation. They were also more likely to report that their family members would feel positively if they supported them with their menstrual health. Surprisingly, non-doers were more likely to report that others would disapprove if they did not support their family.

**Figure 2.**
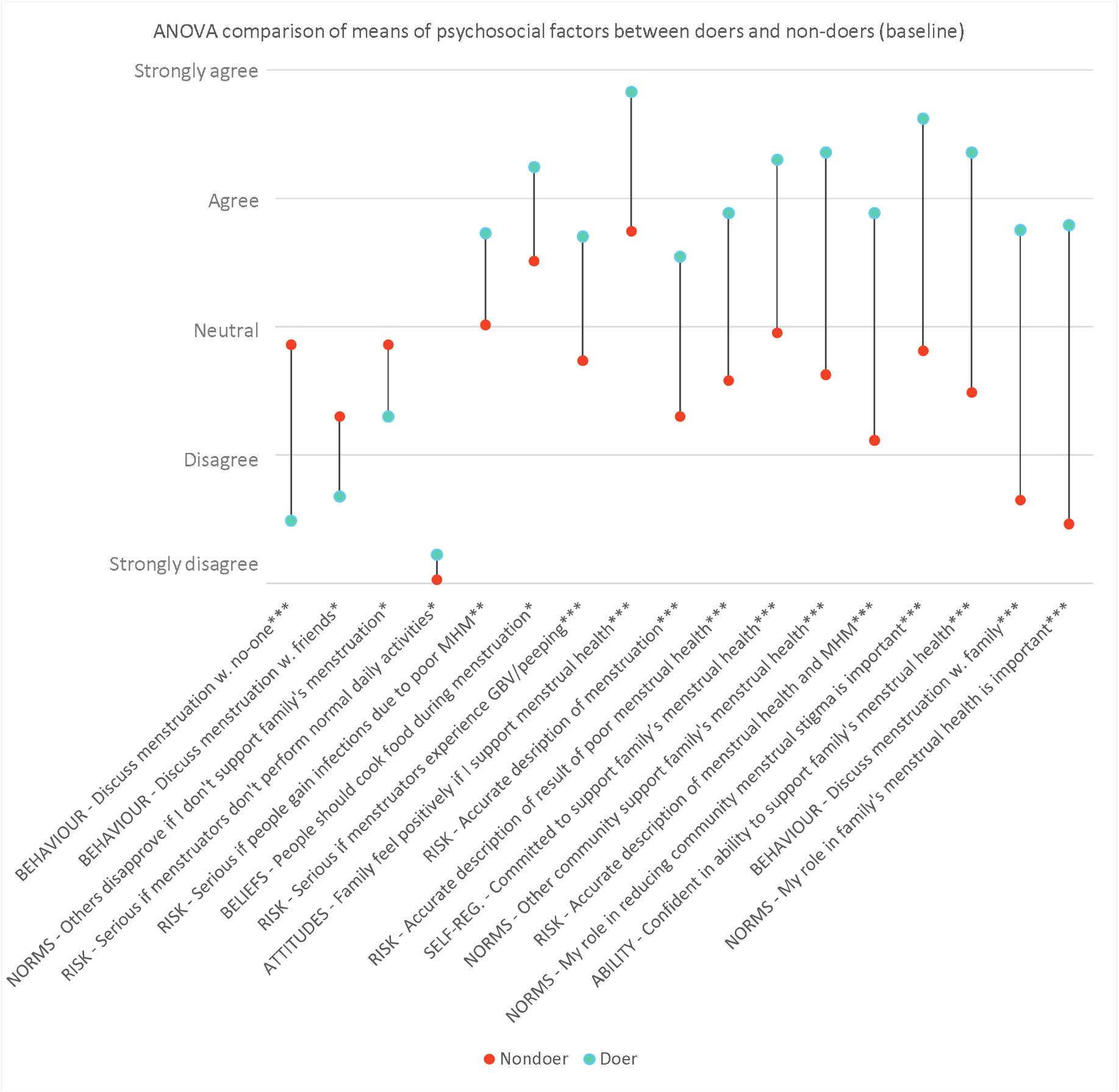
ANOVA comparison of means – doer and non-doer - ^*^p□≤□.05, ^**^p□≤□.01, ^***^p□≤□.001. n = 149

Between the baseline and endline surveys, World Vision implemented their behaviour change programme. The RANAS Behaviour Change Techniques (BCTs) are set to target the specific psychosocial factors that hold significance over the target behaviour. The BCTs that World Vision used are as follows:

| **Psychosocial factors** | **Behaviour change techniques** |
| --- | --- |
|  | *Information BCTs – Risk factors* |
| *Health Knowledge* | BCT 1 Present facts<br>BCT 2 Present scenarios |
| *Vulnerability* | BCT 3 Inform about and assess personal risk |
| *Severity* | BCT 4 Arouse fear |
|  | *Persuasive BCTs – Attitude factors* |
| *Beliefs about Costs and Benefits* | BCT 7 Prompt to talk to others |
| *Feelings* | BCT 8 Describe feelings about performing and about consequences of the behaviour |
|  | *Norm BCTs – Norm factors* |
| *Others' Behaviour* | BCT 9 Inform about others' behaviour |
| *Personal Importance* | BCT 13 Provide a positive group identity<br>BCT 14 Prompt identification as role model |
|  | *Infrastructural, skill and ability BCTs – Ability factors* |
| *How-to-do Knowledge* | BCT 15 Provide instruction |
| *Confidence in Performance* | BCT 17 Demonstrate and model behaviour<br>BCT 20 Facilitate resources |

### Baseline/endline ANOVA analysis

To see if non-menstruator Rohingya attitudes and behaviours towards menstruation changed after World Vision’s intervention, the researchers undertook another ANOVA comparison of means for the overall survey results at baseline and endline. Only responses with a p value ≤_J.05 are shown in **Error! Reference source not found**.. The scales were the same as for the multiple linear regression for RANAS psychosocial factors – yet they have all been put on the same scale from ‘strongly disagree’ to ‘strongly agree’ to make them visually comparable. The results are in order of biggest negative to positive difference between endline and baseline.

**Figure 3.**
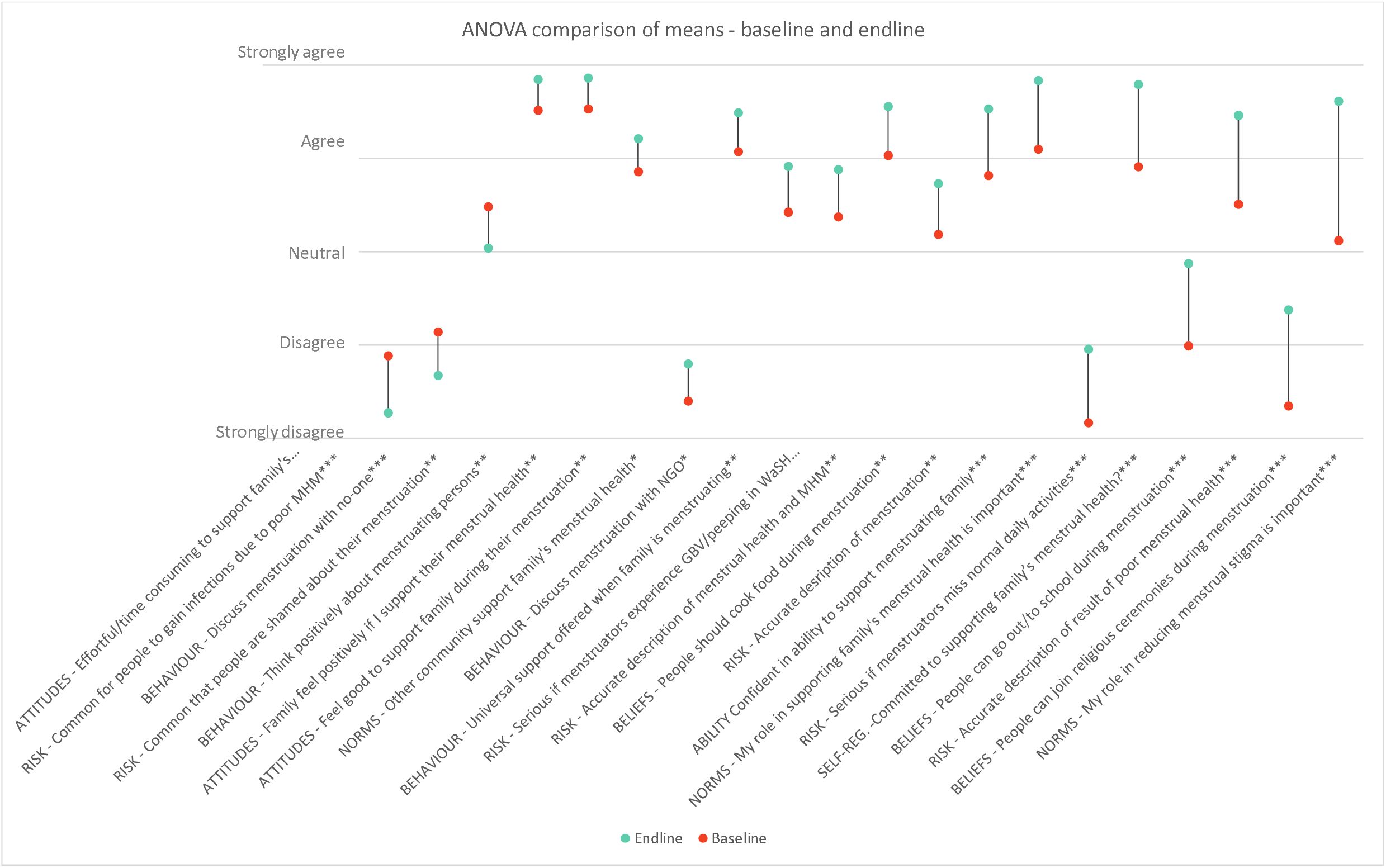
ANOVA comparison of means – baseline and endline - ^*^p□≤□.05, ^**^p□≤□.01, ^***^p□≤□.001. n = 149 at baseline, n= 145 at endline

As we can see, 22 factors changed significantly from baseline to endline. In terms of the three questions that defined the dependent variable – thinking positively about menstruation, supporting family during menstruation, and feeling good to support with menstruation – all improved significantly. The amount of people who discussed menstruation with no one reduced whereas the amount who discuss menstruation with NGOs increased. Perceptions that it is effortful and time consuming to support family with menstruation reduced as did beliefs that it was common for menstruators to be shamed about menstruation and gain infections due to poor MHM. Perceptions that family would feel positively if they were to support them with menstruation and that other community members support their family with menstruation also increased. Beliefs that menstruators should cook food, leave the house, and attend religious ceremonies when menstruating also increased at endline level, along with the reporting of menstruators experiencing GBV in WaSH facilities and not partaking in normal daily activities as a serious matter. Participants at endline gave more accurate descriptions of menstruation, menstrual health, MHM, and the results of poor menstrual health. The endline survey also showed an increase in participants feeling their role in supporting family members and reducing menstrual stigma in the community is important. Confidence and commitment to this also increased.

## Discussion

This section unpacks the regression analyses and doer/non-doer comparison of means to understand how the significant psychosocial and contextual factors influence the desired behaviour. It also examines the psychosocial factors that have changed following World Vision’s programme as revealed by the baseline/endline analysis. The main factors we were interested in seeing change were the ones that defined doers and the dependent variable: Offering universal support when their family members are menstruating, Feeling positive in supporting family members during their menstruation, and Thinking positively about menstruating persons. All of these showed a significant increase from baseline to endline following the intervention.

### Context

Expectedly, there was a relationship between marital status and the desired behaviour. Those who were married were 110% more likely to have positive attitudes and behaviours towards menstruation. Not only does being married to someone who menstruates give non-menstruators the opportunity to practise the desired behaviour, studies on the way non-menstruators either come to learn or become more sympathetic towards menstruation is if/when they have an intimate relationship with someone who menstruates (Allen et al. 2011). This US study showed how non-menstruators reported feeling they had arrived at a place of maturation in their attitudes towards menstruation owed to this empathic, familiar relationship (Allen et al., 2011). Doers were also more likely to have children and be over the age of 25, which coincides with them being married.

In terms of location, it was interesting to see that those living in block A were much more likely to hold negative views and those living in block C and F more likely to have positive views than the other blocks. This shows that even across the same demographics and programme interventions specific, localised issues can make a measureable difference. It is likely due to the levels of conflict experienced in Block A; when safety and security needs are compromised levels of participation in programmes such as these diminish (UNHCR, 2020). It is reassuring that those who had already been engaged by World Vision regarding menstruation were more likely to have the desired attitudes and behaviours showing that their previous efforts before the study were having some effect.

Doers were more likely to have arrived before the 2017 influx and non-doers during, meaning doers may have had more opportunity to be exposed to preceding menstrual health programmes. Non-doers were more likely to have learnt about menstruation age 5-20 and from friends or an Imam/Quran, whereas doers were more likely to have first learned about menstruation age 21-30 and from family and NGOs. This may be that non-doers received uninformed or negative information from friends or an Imam when they were younger, whereas doers received more rigorous and positive explanations from their spouses or World Vision, as is explored in the following paragraphs.

### Who non-menstruators discuss menstruation with

Both the regression analysis and the doer/non-doer comparison of means showed a relationship between not discussing menstruation with anyone and not demonstrating the desired behaviour. If someone doesn’t discuss menstruation with anyone they are unlikely to be able to understand what menstruation is or form sympathetic opinions towards those who experience it. Equally, by harbouring negative or avoidant views towards menstruation, the person may continue not to engage in the subject with anyone, sustaining the cycle. Non-menstruators are frequently either left out of the conversation or choose not to participate in it, perpetuating the notion that menstruation is purely a ‘womens’ issue’ as opposed to a human rights issue (Bhattacharjee 2020; Babbar et al. 2022). In this way, the problem remains unaddressed.

Non-doers were also much more likely to speak to non-menstruator friends about menstruation. Although this may seem positive, research shows that when the information non-menstruators receive is from other non-menstruator friends this can become misconstrued with the sharing of false or misunderstood knowledge (Erchull 2020). Conversely, there was a significant difference in doers being more likely to speak to family about menstruation - a desired outcome of the programme. Studies show that non-menstruators start to learn more about menstruation and change their opinions towards it once they start having conversations with their wives or menstruating family members (Shah et al. 2023). The amount of people reporting that they speak to NGOs about menstruation increased significantly from baseline to endline, along with the people speaking to no-one decreasing, which is tautological since the intervention involved them talking to NGOs about menstruation.

### Beliefs

None of the beliefs studied showed a significant relationship with the desired attitudes and behaviours in the regression analysis, however it was found that doers were more likely to believe that menstruators should still cook food whilst they’re menstruating. From baseline to endline the number of people believing that menstruators should go out, to school, and to religious ceremonies during menstruation increased. Trying to change a community’s customary practises can be paternalistic, elitist and a form of neo-colonialism. There is a balance to be struck between addressing practises that are harmful to a persons’ health, for example Female Genital Mutilation and those that are not, for example not touching cattle whilst menstruating. Though some menstrual practises among the Rohingya may affect menstruator health and wellness, for instance not leaving the house whilst menstruating, World Vision has designed their programme in a way for the community to decide how they wish to shape themselves, which may be to adapt around existing practises rather than change culture.

### Risk

Doers were much more likely to report thinking it was of serious concern if menstruators gain infections due to poor MHM, experience GBV or peeping when using WaSH facilities, or do not perform normal daily activities due to customary practices. Having awareness of the negative effects of poor MHM and grading this as serious may be a motivator for wanting to assist family members to ensure they have the right materials, facilities, and space to manage their periods properly. Expectedly, doers were also more likely to be able to give an accurate description of menstruation, menstrual health and MHM, and the results of poor menstrual health. Studies have shown that non-menstruators who receive comprehensive education about menstrual health are more likely to exhibit supportive attitudes and behaviours towards menstruators (Eyring et al. 2023; Hennegan and Montgomery 2016; Shah et al. 2023). These educational programmes aim to dispel myths and taboos associated with menstruation, thereby fostering a more supportive environment. The ability to give an accurate description of each of these elements of menstruation increased from baseline to endline after World Vision’s intervention.

Regarding GBV or peeping as a serious matter may indicate a level of awareness and empathy towards the negative, gendered experiences menstruators endure, which may extend to other aspects of menstrual health. To reiterate the official definition of menstrual health, this goes beyond access to materials and facilities to encompass being able to manage ones period in a supportive environment that is safe, private, and comfortable. By having an awareness of the GBV and peeping risk menstruators face when accessing WaSH facilities, they may also realise the impact this has on being able to manage ones period.

Agreeing that it’s serious if menstruators experience GBV or peeping or if they’re restricted from normal daily activities increased from baseline to endline, however perception of the severity of them gaining infections due to poor MHM did not. Two factors which didn’t show any significance in the regression or ANOVA analyses but which did have a significant change from baseline to endline was perceiving that it’s common for menstruators to (a) gain infections due to poor MHM and (b) be shamed about their menstruation. This may show that the programme has enhanced the perception of how at risk menstruators are to gaining infections, especially in the environment of a refugee camp, and the regularity to which they face menstrual discrimination. It could even be supposed that non-menstruators were aware of menstrual discrimination beforehand but didn’t think of it as an issue. Now being more acutely aware of it, they may be more likely to see its prevalence.

### Attitudes

Both the regression analysis and ANOVA test between doers and non-doers showed the belief that their family would feel positively if they support them during menstruation had a significant influence on having positive attitudes and behaviours. The Theory of Planned Behaviour coined by Icek Ajzen posits that if individuals perceive that changing their behaviour will have positive consequences for their family and friends, they are more likely to have favourable attitudes toward adopting the behaviour (1987). This factor increased from baseline to endline. A factor that showed a significant decrease from baseline to endline, which did not show any significance in the regression and ANOVA analyses, was reporting that supporting family members was effortful and time-consuming. Perhaps by having a greater understanding of what is required of them and putting this into practise, doers were able to see that supporting their family during menstruation is less taxing than what it might be perceived to be by those not carrying out the behaviours, which may be a factor preventing them from offering such support.

### Norms

Another factor, which showed significance in both analyses, was the individual feeling that their role in supporting their family’s menstrual health was important. Again relating to the Theory of Planned Behaviour, Subjective Norms refer to an individual’s perception of social pressure or expectations from others. If an individual perceives their role within their family or community as important and that others expect or support them to engage in certain behaviours, they are more likely to feel motivated to comply with those norms and adopt the behaviour (Azjen, 1991). Though the main aim of the programme was to encourage supportive behaviours within the home, the doer/non-doer analysis also showed that doers felt their role in reducing stigma among the community was important. Both of these factors increased from baseline to endline. Another factor that also increased was doers perceiving that other non-menstruators in the community were supporting their family during menstruation, which may influence their perception of the social expectations to adopt the behaviour. The presence of non-menstruator role models who actively support menstrual health initiatives can significantly influence other non-menstruators. When community leaders or public figures advocate for menstrual health, it helps normalize the conversation and encourages other non-menstruators to adopt supportive behaviours (Hennegan and Montgomery 2016). For example, this research has shown that religion and Imams are highly influential in this community: when the Imam has a negative view on menstruation, this is echoed in the community and vice versa when the Imam holds positive and supportive views. This research does not intend to criticise Islam in perpetuating negative attitudes but to recognise that religious texts of any kind can be interpreted in such a way among different groups and used to influence the population. Equally, religion and religious figures can also influence very positive attitudes towards menstruation and menstruators.

Non-doers reported feeling that others would disapprove if they didn’t support their family during menstruation. This didn’t show any significant change between baseline and endline. The motivation to comply with others’ approval or disapproval is an assessment of how important it is to have the approval from important others (Peters et al., 2010). This may show that others’ disapproval is not an important psychosocial factor to influence this behaviour in this community.

### Ability

Doers reported feeling more confident in their ability to support their family during menstruation. Perceived Behavioural Control in the Theory of Planned Behaviour emphasizes the role of self-efficacy in shaping behaviour change. If an individual has high self-efficacy, they are more likely to believe they have the skills, knowledge and resources necessary to carry out the behaviour well (Azjen, 1991). Furthermore, they are more likely to believe their efforts will lead to the desired outcomes. This in turn means they are more likely to set challenging goals for themselves and persist even in the face of obstacles or setbacks. Related to the perception of their own role in the community and others’ behaviour, observing others who are similar to oneself successfully performing a behaviour can enhance self-efficacy by providing social proof (Bandura 1994). Confidence in the behaviour increased from baseline to endline.

### Self-regulation

Feeling committed to continuing to support their family’s menstrual health showed a significant relationship with the desired behaviour. This relates to the ‘self-regulation’ aspect of RANAS, which takes into consideration how although behaviour change programmes may seem to be effective initially, over time and without regular prompts, the new behaviour may start to dwindle. There is little research into what sustains non-menstruator engagement in gender-related programmes. In terms of studies on non-menstruator engagement in GBV prevention work there is some evidence to suggest that non-menstruator commitment links to ‘tangible feelings of making a difference… opportunities for discussion and reflection about social justice and GBV, and a sense of membership in a supportive community in which there is permission to do “masculinity” differently’ (Tolman et al., 2019). Sustaining continued non-menstruator participation is one of the primary global challenges to such programmes (Casey et al., 2013). Commitment to the behaviour increased from baseline to endline. However, World Vision was not implementing any BCTs that addressed this factor so we cannot say the positive changes we have seen in this study will withstand over time.

What can be seen in all of these results is an indication of which factors have an influence on having positive attitudes and behaviours towards menstruation. What it lacks is an exploration of how and why. Additionally, the RANAS approach does not provide guidance on how to consider or address contextual factors. We suggest that what is required next is a Realist Evaluation using qualitative interviews with the participants as it would allow us to unpack the mechanisms generating these changes for each significant factor found in this study.

### Limitations and positionality

Working between languages always presents opportunity for meaning to get lost in translation. There may have been instances where the data collectors and/or participants inferred a different meaning from the survey questions than what was meant by the survey creators. Additionally Likert scales may not be well understood or naturally adopted as a means of measurement. To get around this, the researchers often created a scale retrospectively based on participant responses when the original scales were not followed.

Due to time restrictions in access to the camp, the lead author was only able to conduct the first 10 surveys alongside the data collectors. This meant she was not available to detect when the survey question may have been misunderstood or to answer any clarifying questions from data collector or participant. The sample size of 150 participants is not necessarily generalizable to the one million Rohingya living in Bangladesh.

Positionality statements have been criticised as being a form of neo-colonialism wherein White and/or Western researchers are either still asserting dominance through the statement of assumed hierarchies or giving lip-service to recognising and undoing bias without being truly reflexive (Gani and Khan 2024). Nonetheless, we include one in order to provide more clarity on our position for the reader. The lead author is White-British, Middle-Class, and non-religious. She recognises that much of her privilege to be funded and able to conduct research in the international sphere stems from the colonial legacies of the British Empire. She understands that behaviour change programmes can be neo-colonial and that she is limited when comprehending the nuances of the religious and cultural characteristics of the population studied in this paper. The three research partners from UNICEF and World Vision who are authors to this paper are Bangladeshi and have been working closely with the Rohingya population since their arrival to Cox’s Bazar in 2017. Though possessing many cultural, religious and, linguistic similarities to the Rohingya, which has advised the research, they are still outsiders to this population so will possess their own misunderstandings and biases.

## Conclusion

This paper has used a case study in Kutupalong camp with Rohingya refugees to identify psychosocial and contextual factors that are determinants of positive attitudes and behaviours towards menstruation and menstruators among those that don’t menstruate – namely cisgender men and boys. The study offered an exceptional example of a strongly patriarchal population living within the challenging environment of a refugee camp. Nonetheless, the programme saw changes in perceptions of menstruation and supportive behaviours among the population. Whether these changes will be sustained is another matter.

The influential psychosocial factors included who participants discussed menstruation with, beliefs about menstruation, the perceived severity of menstrual health issues, family reaction, menstrual knowledge, commitment and confidence. Influential contextual factors included house location, marital status, having children, age, arrival year in camp, previous menstrual education from World Vision, and how and at what age they were introduced to menstruation.

ANOVA comparisons showed positive changes in 22 variables from baseline to endline following the intervention. Participants increasingly thought positively about menstruation, supported family members, and felt good about providing support. Discussions with NGO workers increased, while views on upholding customary menstrual norms decreased. Participants at endline perceived less effort in supporting family, noted a decrease in perceived commonality of menstrual health issues, and gave more accurate descriptions of menstrual health. They felt their role in supporting family and reducing menstrual stigma was important, with increased confidence and commitment to these behaviours.

It is imperative to engage non-menstruators in these programmes if we are to see sustained improvements to menstrual health. Historically we have only addressed physical needs of facilities and materials, or improving education and reducing stigma in the half of the population who menstruate. This is especially important in patriarchal populations where the social structures and menstrual stigma give less power to those who menstruate in ameliorating the situation. This papers suggests which contextual and psychosocial factors are likely the most beneficial to target in a behaviour change campaign to improve menstrual attitudes and behaviours within this context. Through this, many varied barriers to menstrual health that menstruators face in this setting may be reduced and a more supportive environment created. Accordingly, overall health can be improved, dignity protected, and equity between those who do and do not menstruate enhanced.

The same research methods used in this paper can be used to discover factors to be targeted in other populations and contexts. Though the RANAS approach used in this paper provides important insights on the psychosocial factors that influence behaviours towards menstruation, what it lacks is an exploration of how to address contextual factors. Being largely based on quantitative data, it also does not offer a method for unpacking the generative causes for these factors to bring about change. Such a method would be Realist Evaluation, using theorised qualitative interviews with programme stakeholders participants.

## Data Availability

All data produced in the present study are available upon reasonable request to the authors

## Supplementary material

Factors with a correlation of <20% excluded from contextual regression analysis

a. Lives in Block B, D or E
b. Educational Level of self or mother or father
c. Arrived after 2017 influx
d. Learnt about menstruation age 10-15 or 30+
e. First learnt about menstruation from friends or institution/teacher/book or unsure
f. Learnt more about menstruation from friends or institution/teacher/book or Imam/Quran or NGO or unsure

Factors with a correlation of <20% excluded from psychosocial regression analysis

a. Thinks girls/women should go out/to school or join religious ceremonies or cook food during menstruation
b. Discusses menstruation with friends or NGO or Imam/teacher
c. Acceptable for a non-menstruator to use a ‘female’ toilet or shower block
d. Common for women to experience GBV or peeping when using WaSH facilities or are shamed or teased about their menstruation or don’t participate in normal daily activities as per customary practise
e. Serious if women don’t participate in normal daily activities as per customary practise
f. Expensive to buy menstrual materials for family members
g. Very effortful and time consuming to support family members with menstruation

## References

Ajzen I. The Theory of Planned Behavior. Organizational Behavior and Human Decision Processes. 1991;50(2):179– 211.

Ajzen, I. Attitudes, Traits, and Actions: Dispositional Prediction of Behavior in Personality and Social Psychology, Editor(s): Leonard Berkowitz, Advances in Experimental Social Psychology, Academic Press, Volume 20, 1987, Pages 1–63.

Allen, K. R., Kaestle, C. E., & Goldberg, A. E. (2011). More Than Just a Punctuation Mark: How Boys and Young Men Learn About Menstruation. Journal of Family Issues, 32(2), 129–156. 10.1177/0192513X10371609

Babbar K. Martin J. Ruiz J. Ahmad Parray A. Sommer M. 2022. Menstrual health is a public health and human rights issue. The Lancet. VOLUME 7, ISSUE 1, E10–E11.

Bandura, A. (1994). Self-efficacy. In V.S. Ramachaudran (Ed.), Encyclopedia of human behavior (Vol. 4, pp. 71-81). New York: Academic Press. (Reprinted in H. Friedman [Ed.], Encyclopedia of mental health. San Diego: Academic Press, 1998).

Bhattacharjee, M. 2020. Menstruation inJEmergencies: Developing aJPeriod-Friendly Emergency Response in The Palgrave Handbook of Critical Menstruation Studies. ISBN 978-981-15-0613-0 ISBN 978-981-15-0614-7 (eBook) 10.1007/978-981-15-0614-7

Casey EA, Carlson J, Fraguela-Rios C, Kimball E, Neugut TB, Tolman RM, Edleson JL. Context, Challenges, and Tensions in Global Efforts to Engage Men in the Prevention of Violence against Women: An Ecological Analysis. Men Masc. 2013 Jun;16(2):228–251. doi: 10.1177/1097184X12472336. PMID: 25568612; PMCID: PMC4283930.

Day, I. 2024. Menstruation, Human Rights and the Patriarchy: How International Human Rights Law Puts Menstruating People at Risk. Harvard Human Rights Journal. 37.

Erchull, M. 2020. “You Will Find Out When the Time Is Right”: Boys, Men, and Menstruation in The Palgrave Handbook of Critical Menstruation Studies. ISBN 978-981-15-0613-0 ISBN 978-981-15-0614-7 (eBook) 10.1007/978-981-15-0614-7

Eyring, J.B., Crandall, A. & Magnusson, B.M. A Modified Menstrual Attitudes Scale: Heteronormative Attitudes, Sexism, and Attitudes Toward Menstruation in Male and Female Adults. Arch Sex Behav 52, 1535–1547 (2023). 10.1007/s10508-023-02565-y

Gani, J.K. and Khan, R.M., 2024. Positionality statements as a function of coloniality: Interrogating reflexive methodologies. International Studies Quarterly, 68(2), p.sqae038.

Glass N, Perrin N, Marsh M, Clough A, Desgroppes A, Kaburu F, Ross B, Read-Hamilton S. Effectiveness of the Communities Care programme on change in social norms associated with gender-based violence (GBV) with residents in intervention compared with control districts in Mogadishu, Somalia. BMJ Open. 2019 Mar 13;9(3):e023819. doi: 10.1136/bmjopen-2018-023819. PMID: 30872541; PMCID: PMC6429733.

Hennegan J, Montgomery P (2016) Do Menstrual Hygiene Management Interventions Improve Education and Psychosocial Outcomes for Women and Girls in Low and Middle Income Countries? A Systematic Review. PLoS ONE 11(2): e0146985. 10.1371/journal.pone.0146985

Pandit K, Hasan MJ, Islam T, Rakib TM. Constraints and current practices of menstrual hygiene among Rohingya adolescent girls. Heliyon. 2022 May 19;8(5):e09465. doi: 10.1016/j.heliyon.2022.e09465. PMID: 35647348; PMCID: PMC9136249.

Patel Kripalini, Panda Nishisipa, Sahoo Krushna Chandra, Saxena Shipra, Chouhan Narendra Singh, Singh Pratibha, Ghosh Upasona, Panda Bhuputra. 2022. A systematic review of menstrual hygiene management (MHM) during humanitarian crises and/or emergencies in low- and middle-income countries. Frontiers in Public Health. 10.

Peters RM, Templin TN. Theory of planned behavior, self-care motivation, and blood pressure self-care. Res Theory Nurs Pract. 2010;24(3):172–86. doi: 10.1891/1541-6577.24.3.172. PMID: 20949834; PMCID: PMC3728772.

Power and Sample size. 2024. [Online] Available from: http://powerandsamplesize.com/Calculators/Compare-2-Means/2-Sample-1-Sided. [Accessed 12 August 2024].

Rahaman, K.S., Ramos, S., Harter, M., Mosler, JH. 2022. Psychosocial factors influencing handwashing behaviour and the design of behaviour change interventions for the Rohingya camps in Bangladesh. Journal of Water, Sanitation and Hygiene for Development. 12 (10): 671–682. doi: 10.2166/washdev.2022.167

Ranas (2023). A practical guide using the RANAS approach to systematic behaviour change. Zurich, Switzerland.

Ranas Ltd. (2022). The RANAS approach to systematic behavior change. Methodological Fact Sheet 1. Zürich, Switzerland.

Rohwerder. 2014. https://assets.publishing.service.gov.uk/media/57a089a6e5274a27b20001cf/hdq1107.pdf

Shah SF, Punjani NS, Rizvi SN, Sheikh SS, Jan R. Knowledge, Attitudes, and Practices Regarding Menstrual Hygiene among Girls in Ghizer, Gilgit, Pakistan. International Journal of Environmental Research and Public Health. 2023; 20(14):6424. 10.3390/ijerph20146424

Sommer, M., Schmitt, M., Clatworthy, D. (2017). A toolkit for integrating Menstrual Hygiene Management (MHM) into humanitarian response. (First edit). New York: Columbia University, Mailman School of Public Health and International Rescue Committee.

Sommer, Marni, Margaret L. Schmitt, David Clatworthy, Gina, Bramucci, Erin Wheeler, and Ruwan Ratnayake. 2016. “What Is the Scope for Addressing Menstrual Hygiene Management in Complex Humanitarian Emergencies? A Global Review.” Waterlines, Practical Action Publishing 35 (3): 245–64. 10.3362/1756-3488.2016.024.

Sphere Project, Sphere Handbook 2018. 2018. Available from: https://spherestandards.org/handbook-2018/

Tellier M. Alex Farley, Andisheh Jahangir, Shamirah Nakalema, Diana Nalunga, and Siri Tellier. 2020. Chapter 45Practice Note: Menstrual Health Management in Humanitarian Settings In The Palgrave Handbook of Critical Menstruation Studies.

Tolman, R. M., Casey, E. A., Allen, C. T., Carlson, J., Leek, C., & Storer, H. L. (2019). A Global Exploratory Analysis of Men Participating in Gender-Based Violence Prevention. Journal of Interpersonal Violence, 34(16), 3438–3465. 10.1177/0886260516670181

UNHCR. Tearing down the walls: Confronting the barriers to internally displaced women and girls’ participation in humanitarian settings. UNHCR; 2020. [Online]. Available at https://www.unhcr.org/uk/media/tearing-down-walls-confronting-barriers-internally-displaced-women-and-girls-participation. Accessed 20 Apr 2021.

Winkler IT, Lhaki P, Baumann SE. Its manifestations may vary, but menstrual stigma is universal. Women’s Health. 2024;20.

